# Public perspectives towards extended community pharmacies services in Saudi Arabia: an online cross-sectional study

**DOI:** 10.1101/2022.12.21.22283819

**Authors:** Khalid S. Alghamdi, Max Petzold, Ashraf A. Ewis, Mahdi H. Alsugoor, Khalid Saaban, Laith Hussain-Alkhateeb

## Abstract

**Background:** In many developed countries, the scope of community pharmacy services has extended to deliver more advanced applications. Unlike the traditional practice focusing on pharmaceutical sales, extended community pharmacy services (ECPSs) are patient-centred, typically offered by specialised healthcare centres, which improve public health, reduce pressure imposed on healthcare professionals and rationalise the health system expenditure. However, several studies have revealed that community pharmacies (CPs) provide only marginalised services. Therefore, a public review is crucial for effectively utilising such healthcare-complementary professional services. This study explores CPs utilisation among the Saudi public in terms of knowledge, attitudes, and barriers towards ECPSs.

**Materials and Methods:** A cross-sectional web-based survey of a non-probability sample was conducted between October and December 2021. Numerical and graphical descriptive statistics were employed with an additional analytical assessment using binary logistic regression for the association between characteristics of participants and barriers to utilising ECPSs.

**Results:** A total of 563 subjects participated, of which about 33% considered CPs the first place to visit if they have medication concerns. Most were unfamiliar with medication therapy management and health screening services (77%%, and 68%, respectively). Pharmacy clinics for receiving private counselling and electronic patients’ medical records were unknown facilities to them (79% and 63%, respectively). A substantial portion considered the lack of privacy (58%) and inadequate communication with the community pharmacists (56%) as key barriers. The logistic regression analysis revealed that the underdeveloped infrastructure of CPs was significantly associated with almost all factors. Privacy issues concerned females, highly educated and high-income individuals, and those with health insurance. In addition, low socio-economic status, subjects with special needs or chronic diseases, non-health insured and smokers were less likely to utilise the ECPSs due to economic reasons.

**Conclusions:** The most public was under-utilised services and facilities. Positive attitudes were associated with public concerns regarding their privacy and the cost of services. In line with the Saudi vision 2030, supporting CPs and increasing public awareness towards ECPSs will have significant public health implications.

## Introduction

Pharmaceutical care can directly and comprehensively address public health needs. It is “the pharmacist’s contribution to the care of individuals in order to optimise medicines use and improve health outcomes” [1]. Through this approach, community pharmacies (CPs) can assist other healthcare professionals in providing more adequate services focusing on patient-centred care [2]. These CPs services have been categorised by the United Kingdom National Health Services (NHS) into “essential” and “advanced” [3, 4]. Essential services focus on medication dispensing and public health promotion, while advanced services are of extended community pharmacy services (ECPSs) nature provided in addition to the routine essential services [5]. Furthermore, ECPSs may be applied via CPs to carry out health services usually provided by other healthcare professionals (e.g., general practitioners and nurses) [6]. These services may include vaccinations for seasonal flu, a smoking cessation program, health screening (e.g., blood pressure level, blood glucose level, lipids level and bodyweight measurement), medicines use review (medication therapy management) and others[4]. However, CPs’ infrastructure demands well-equipped facilities to ensure efficient service delivery. In the context of this study, the CPs facility refers to any equipment that professionally facilitates CPs practice. These facilities may include automated medication dispensing systems, electronic prescription systems, and medical records systems. In addition, the use of health screening devices in pharmacy clinics for private regular-documented follow-ups.

Based on published reports, mortality and morbidity rates caused by chronic non-communicable diseases can be reduced by early detection through health screening and regular follow-up within CPs [7-9]. Therefore, ECPSs are a more comprehensive approach which aims to improve public health outcomes, reduce the burden on other primary care settings and rationalise the health system expenditure [10]. Nevertheless, the pharmaceutical care framework assumes a successful patient-pharmacist partnership [11]. This fundamental framework is built on caring, trust, good communication, and coordination [12]. Furthermore, patients with long-term health conditions, which entails regular presentation at the healthcare management, are more amenable to receiving care through CPs than seeking physicians’ consultation, primarily due to accessibility and ease of use [13].

Despite the vast array of ECPSs, these services remain under-utilised by many patients who merely rely on essential services. Several studies call for further assessment of public knowledge of ECPSs, which are minimally provided in some countries [14]. For example, the pharmaceutical care concept in the Middle East still needs to be better defined, as well as facing challenges when implementing ECPSs [15]. In Saudi Arabia (SA), pharmacy is considered a professional career pathway despite mounting evidence suggesting that CPs are not well equipped to deliver such services [16]. Nevertheless, research on the general public view about CPs’ role remains a gap in SA [17], which hinders effective public health policymaking. Therefore, this study aims to explore the perceptions of the public as a key stakeholder in this overall healthcare service process, including patients and users of CPs in SA, with an emphasis on; a) identifying patterns of CPs utilisation, b) describing knowledge, attitudes, and the degree of importance towards selected ECPSs and CPs’ facilities, and c) determining any perceived barriers, which could prevent them from attaining ECPSs.

## Materials and Methods

This is a cross-sectional survey using an online self-administered questionnaire conducted in SA. Permission for conducting the study was obtained from the Ethical Research Committee at the Saudi Ministry of Health (MOH), Riyadh (Central IRB log No: 21-56E, June 2021).

### Survey instrument

A literature search was performed to identify research articles with relevant objectives to this study. A modified questionnaire was developed using pre-validated survey items to correspond adequately with the Saudi context [6, 9, 14, 18-20]. The ECPSs included in this study were identified in light of a previously published international systematic review of expanded practice in CPs [21].

Three local staff with multidiscipline within public health, pharmacy, and nursing revised the face and content of the survey for validity. The final version was translated into Arabic by an expert translator using forward-backwards methods until an agreement was reached. The questionnaire was pilot tested on a convenience sample of 33 subjects who were not part of the main study to ensure all questions were explicit. Their feedback was considered for required amendments before the final distribution. Internal consistency was measured by Cronbach’s alpha test, which resulted in an acceptable reliability coefficient of 0.78 (non-standardised items).

The electronic survey included a brief introduction to the study’s aim and informed consent before participating. Respondents were accordingly asked for permission to use their responses anonymously and confidentially. The survey comprised the following five domains, including 18 closed-ended items: a) characteristics of participants, b) patterns of CPs utilisation, c) knowledge of ECPSs and CPs’ facilities, including the degree of importance, d) attitudes towards ECPSs, and e) barriers to using ECPSs (S1 File).

### Sample recruiting and study setting

The total Saudi population is around 34 million inhabitants living across 13 provinces, according to the 2019 Saudi General Authority for Statistics [22]. Therefore, the non-probability convenience sampling technique was adopted based on a sample size estimate generated by the online calculator (Raosoft Inc.) [23].

At least 385 participants were needed to maintain a 95% confidence interval with a 5% margin of error for a proportion of 50%. A larger sample size was targeted at the population of Al Baha, Makkah, and Najran provinces, to account for any missing information that can be expected when using such online tools. All individuals who speak Arabic, are over 18 years old, and use CPs were included in this study.

By the COVID-19 pandemic restrictions imposed in 2021, an online approach utilising the SurveyMonkey platform was used. An electronic link to the survey was distributed via the WhatsApp messenger application, and a snowball recruitment technique was used over three months, starting from the 1^st^ of October until the end of December 2021.

### Data management and statistical analysis

All collected data were cleaned, coded, and analysed using STATA, version 17. Numerical and graphical descriptive statistics were employed. Frequencies and percentages were used to illustrate characteristics of participants, patterns of CPs utilisation, knowledge of ECPSs and CPs’ facilities, and barriers to using ECPSs. In contrast, the degree of importance and attitudes were measured using the 5-Likert scale and then collapsed into the 3-Likert scale for the analysis to describe the public’s responses more clearly. Furthermore, the degree of importance (extremely important/ important = 3, neutral = 2, and not important at all/ not important =1), while the public attitudes (strongly agree/agree = 3, neutral = 2, and strongly disagree/disagree =1).

Additional analytical assessment using logistic regression to explore the association between characteristics of participants and barriers to utilising services. Furthermore, barriers were measured in percentages and later transformed into binary outcomes to fit the probability estimates by logistic regression. For instance, respondents who selected the lack of a pharmacy clinic as a barrier converted to (1) and those who did not select it were assigned null (0). Crude and adjusted logistic regression modelling and the likelihood ratio test of a forward-wise model-building approach were used, and all tests maintained ≤ 5% as a significant level [24]. All potential confounders from the background and characteristics have been assessed in the model.

## Results

### Characteristics of participants

A total of 563 participants consented and were accordingly included in this study. Among them, 68% were males, and 96% were Saudi citizens. Married respondents constituted the majority (66%), and the age group 36 – 45 years old represented the majority (32%) compared to other groups. More than half of the study sample hold a university degree (55%), while 64% were employed, and 52% earned ≤ 12000 in Saudi Riyal per month. Regarding health conditions, 2% of the participants had special needs, 26% were current or previous smokers, and 31% had at least one chronic disease, with only 26 % possessing health insurance. Characteristics of participants are described in Table 1.

**Table 1.** Descriptive statistics of characteristics of participants in Saudi Arabia, N = 563.

| Variables | Frequency | (%) |
| --- | --- | --- |
| **Gender** |  |  |
| Male | 384 | (68.21) |
| Female | 179 | (31.79) |
| **Citizenship** |  |  |
| Saudi | 538 | (95.56) |
| Non-Saudi | 25 | (4.44) |
| **Marital status** |  |  |
| Married | 374 | (66.43) |
| Single and other * | 189 | (33.57) |
| **Age group** |  |  |
| 18 – 25 | 124 | (22.02) |
| 26 – 35 | 133 | (23.62) |
| 36 – 45 | 178 | (31.62) |
| ≥ 46 * | 128 | (22.74) |
| **Educational level** |  |  |
| ≤ High school and Diploma * | 136 | (24.15) |
| University degree | 311 | (55.25) |
| Postgraduate degree (MSc and PhD) * | 116 | (20.60) |
| **Job status** |  |  |
| Employed | 361 | (64.12) |
| Unemployed and Retired * | 202 | (35.88) |
| **Monthly income in SR** |  |  |
| ≤ 12000 * | 291 | (51.69) |
| 12001 – 18000 * | 134 | (23.80) |
| > 18001 * | 82 | (14.56) |
| Undisclosed | 56 | (9.95) |
| **Health insurance** |  |  |
| Yes | 145 | (25.75) |
| No | 418 | (74.25) |
| **Special need** |  |  |
| Yes | 9 | (1.60) |
| No | 554 | (98.40) |
| **Smoking status** |  |  |
| Previous and current smokers * | 149 | (26.47) |
| Non-smoker | 414 | (73.53) |
| **Associated chronic diseases** |  |  |
| Yes * | 172 | (30.55) |
| No | 391 | (69.45) |

\* Groups were collapsed
Abbreviations: SR: Saudi Riyal, MSc: Master of Science, PhD: Doctor of Philosophy

### Public patterns of CPs utilisation

All study participants have been asked about the first place they visit to seek health advice when a minor ailment arises (e.g., headaches, dyspepsia, acute cough, or influenza). Results revealed that the Primary Health Care Centre (PHCC) was the first place to get health advice (29%), followed by CPs (27%).

Also, they have been asked about the first place to visit for any medication concerns (e.g., doubts about dosage, indications, adverse effects, or how it is used). One-third of the participants considered CPs the first place to visit (33%). Interestingly, 19 % of them searched online medical websites for the same reason, while 16% visited PHCC.

Participants then reported one or more purposes of their visits to CPs during the last year. Nearly 54% was mainly for purchasing sanitary and cosmetic products, 53% for collecting prescription only medicine (POM) compared to 49% for buying over the counter medication (OTC), and only 12% for seeking other healthcare services. See Fig 1.

**Fig 1.**
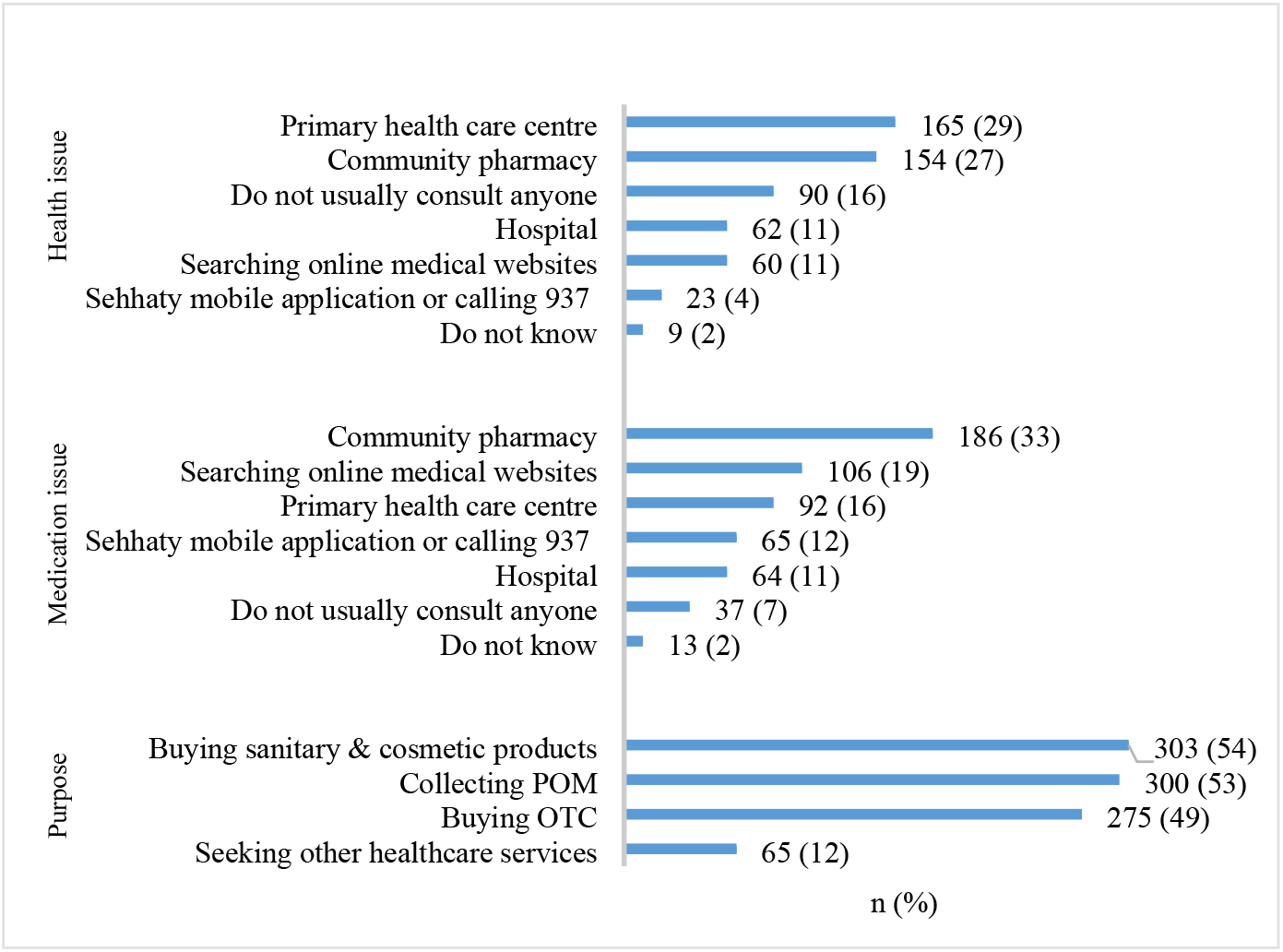
Public patterns of community pharmacies utilisation in Saudi Arabia, N =563.

### Public knowledge of ECPSs and CPs’ facilities

Out of 563 respondents, 540 (96%) stated to what extent they are knowledgeable about ECPSs and CPs’ facilities. However, most respondents do not know whether ECPSs exist or utilise facilities. Amongst selected services, travel health program was known to only (21%), medication therapy management (23%), health screening (32%), health education and promotion program (38%), and smoking cessation program (48%). In comparison, the immunisation service was quite known by most respondents (71%).

Regarding CPs’ facilities, the pharmacy clinic to receive private counselling in a designed closed area and electronic patients’ medical records were known to only a few respondents (21%, and 36%, respectively). In contrast, an electronic prescription system (e.g., Wasfaty: a national prescription system linked to CPs) and online pharmaceutical care, including prescription delivery, were quite known by half of the respondents (54% and 50%, respectively). See Fig 2.

**Fig 2.**
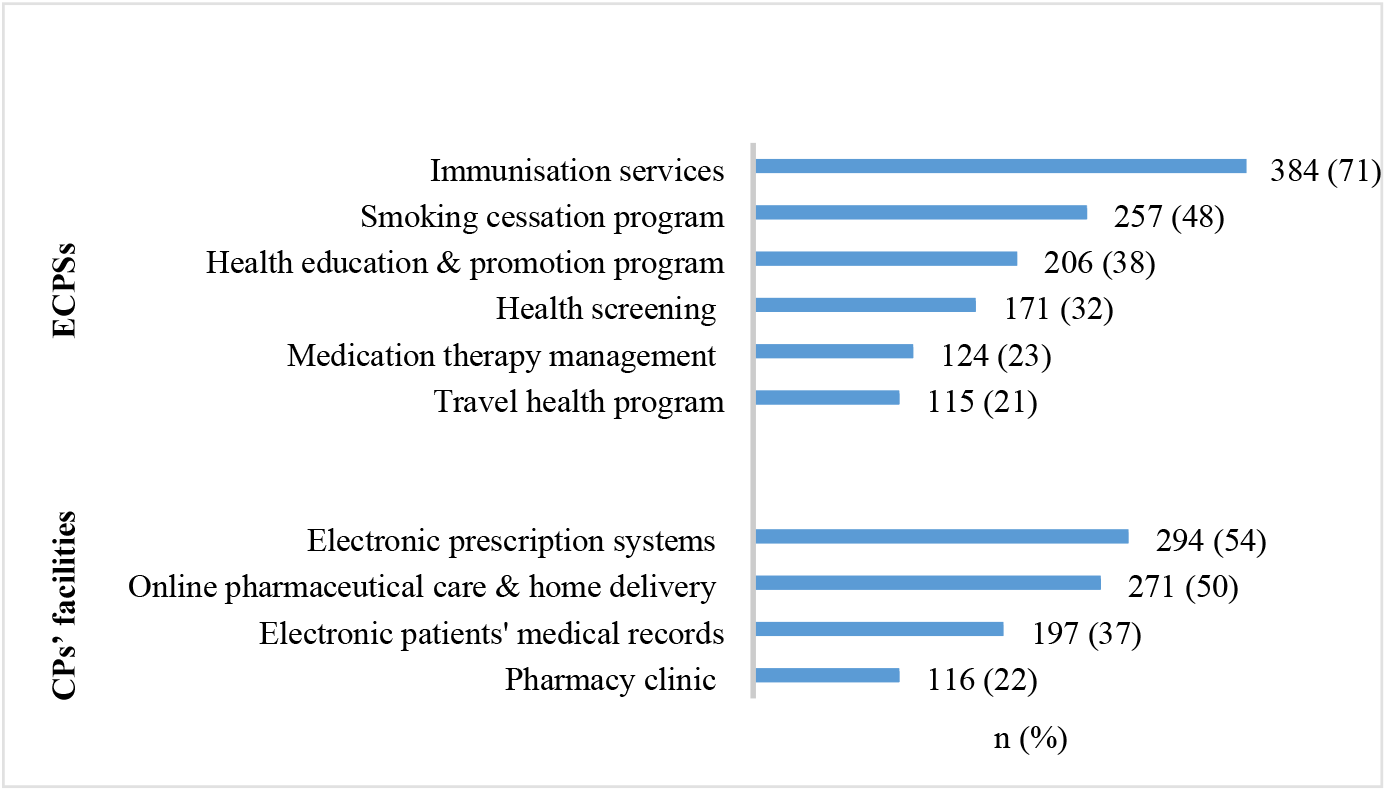
Public knowledge of extended community pharmacies services and facilities in Saudi Arabia, N = 540.

### The degree of importance of ECPSs and CPs’ facilities

Out of 563 participants, 540 (96%) rated the degree of importance of these services and facilities at a critically high level. Providing immunisation services as ECPSs and utilising electronic patients’ medical records as a facility were the most important according to the public (88% and 86%, respectively). More details are presented in Table 2.

**Table 2.**
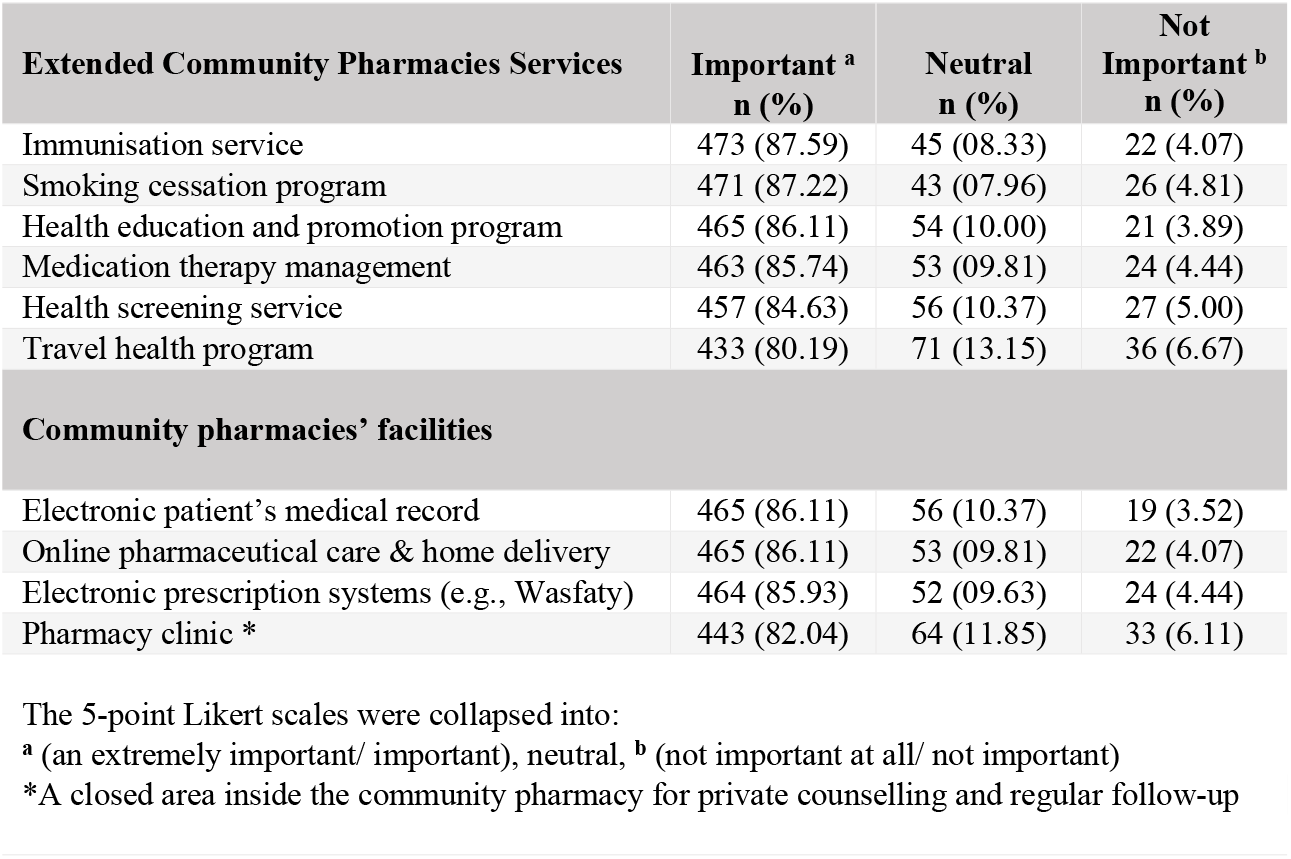
The degree of importance of ECPSs and CPs’ facilities by public of Saudi Arabia, N = 540.

### Public attitudes towards ECPSs

Out of 563 participants, 535 (95%) stated their attitudes. About 84% recommended implementing ECPSs, of which 71% thought that CPs could offer similar services to other healthcare centres, and 66% trusted that community pharmacists are adequately qualified to deliver ECPSs. However, if some of these services are chargeable, 31% were willing to pay, while 35 % were in a neutral position. Table 3 shows more details about public attitudes.

**Table 3.** Public attitudes towards ECPSs in Saudi Arabia, N = 535.

| Statements | Agree <sup>a</sup><br>n (%) | Neutral<br>n (%) | Disagree <sup>b</sup><br>n (%) |
| --- | --- | --- | --- |
| I recommend the ECPSs implementation | 447 (83.55) | 66 (12.34) | 22 (4.11) |
| CPs could provide ECPSs like other clinics | 379 (70.84) | 101 (18.88) | 55 (10.28) |
| If any ECPS is implemented, I will register | 378 (70.65) | 132 (24.67) | 25 (4.67) |
| Pharmacists are qualified to provide ECPSs | 351 (65.61) | 134 (25.05) | 50 (9.35) |
| If some ECPSs are paid the price, I will pay | 167 (31.21) | 187 (34.95) | 181 (33.83) |
| Current traditional services in CPs are enough | 145 (27.10) | 147 (27.48) | 243 (45.42) |

The 5-point Likert scales were collapsed into:
<sup>a</sup> (strongly agree/agree), neutral, <sup>b</sup> (strongly disagree/disagree)
ECPSs: Extended Community Pharmacies Services, CPs: Community Pharmacies

### Barriers to using ECPSs

Out of 563 participants, 533 (95%) selected at least one or more barriers that may prevent them from using ECPSs. More than half of respondents considered the lack of a pharmacy clinic inside CPs to receive private counselling a key barrier (58%). Another half attributed it to the inability to communicate adequately with the community pharmacists (56%). At the same time, other participants saw the lack of interprofessional connection between physicians and community pharmacists to discuss their health conditions as a barrier (45%). In addition, nearly half of CPs users believed that the underdeveloped infrastructure of CPs and staff shortages are barriers to providing these services (47%). The economic status of patients was another barrier (44%), while only 27 % considered the lack of trust in community pharmacists as a barrier. Fig. 3 illuminates barriers arranged to form the highest rated to the lowest.

**Fig 3.**
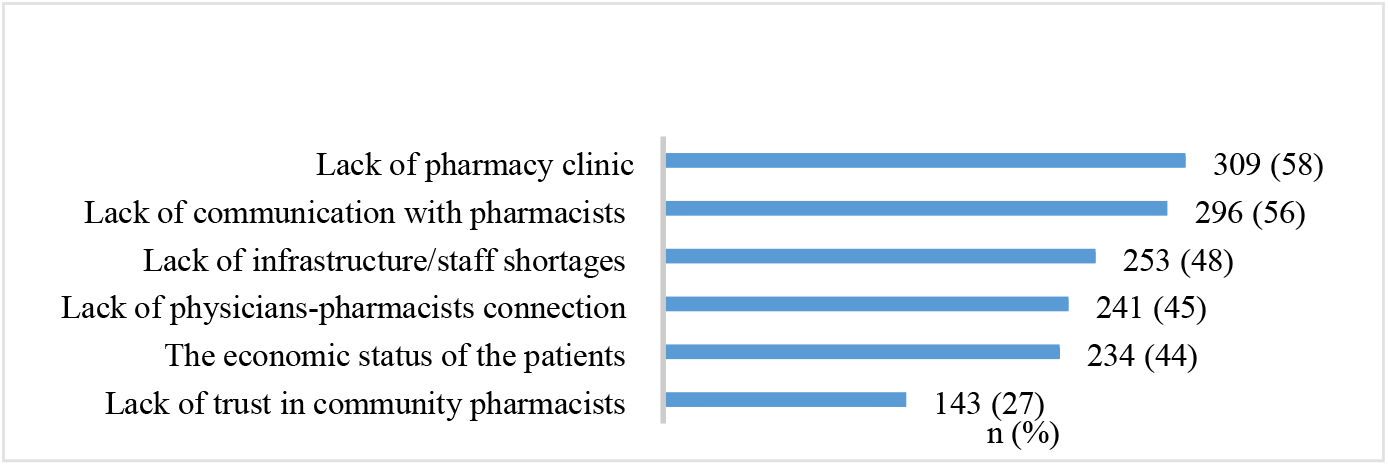
Barriers to using extended community pharmacies services in Saudi Arabia, N = 533.

According to the logistic regression analysis, four barriers were statistically associated with multiple backgrounds and characteristics of the respondents. No significant association was observed for the lack of physicians-community pharmacists connection and patients-community pharmacist communication.

The underdeveloped CPs’ infrastructure, including staff shortage was significantly associated with almost all factors, while the lack of pharmacy clinics was significantly associated with age groups and job status. Moreover, females, individuals with high educational levels and incomes, with health insurance, nonchronic disease and previous and current smokers were more likely to be concerned about their privacy in the lack of pharmacy clinics.

The lack of trust in community pharmacists was associated with gender, monthly income, smoking habit, and first places to visit asking about health or medication issues. Furthermore, married individuals, age groups between 26 – 45 years old, individuals with ≥ a university degree, with health insurance and chronic disease, and with no special needs were less likely to trust community pharmacists.

The patient’s economic status was significantly associated with the respondent’s marital status and smoking habit. Furthermore, males, age groups ≥ 26 years old, individuals with ≤ diploma degree, 12001 – 18000 monthly incomes, non-health insured, special needs, and chronic diseases were more likely to consider the cost of ECPSs.

Table 4 illustrates the analytical assessment of barriers to using ECPSs by different background and characteristics factors.

**Table 4.**
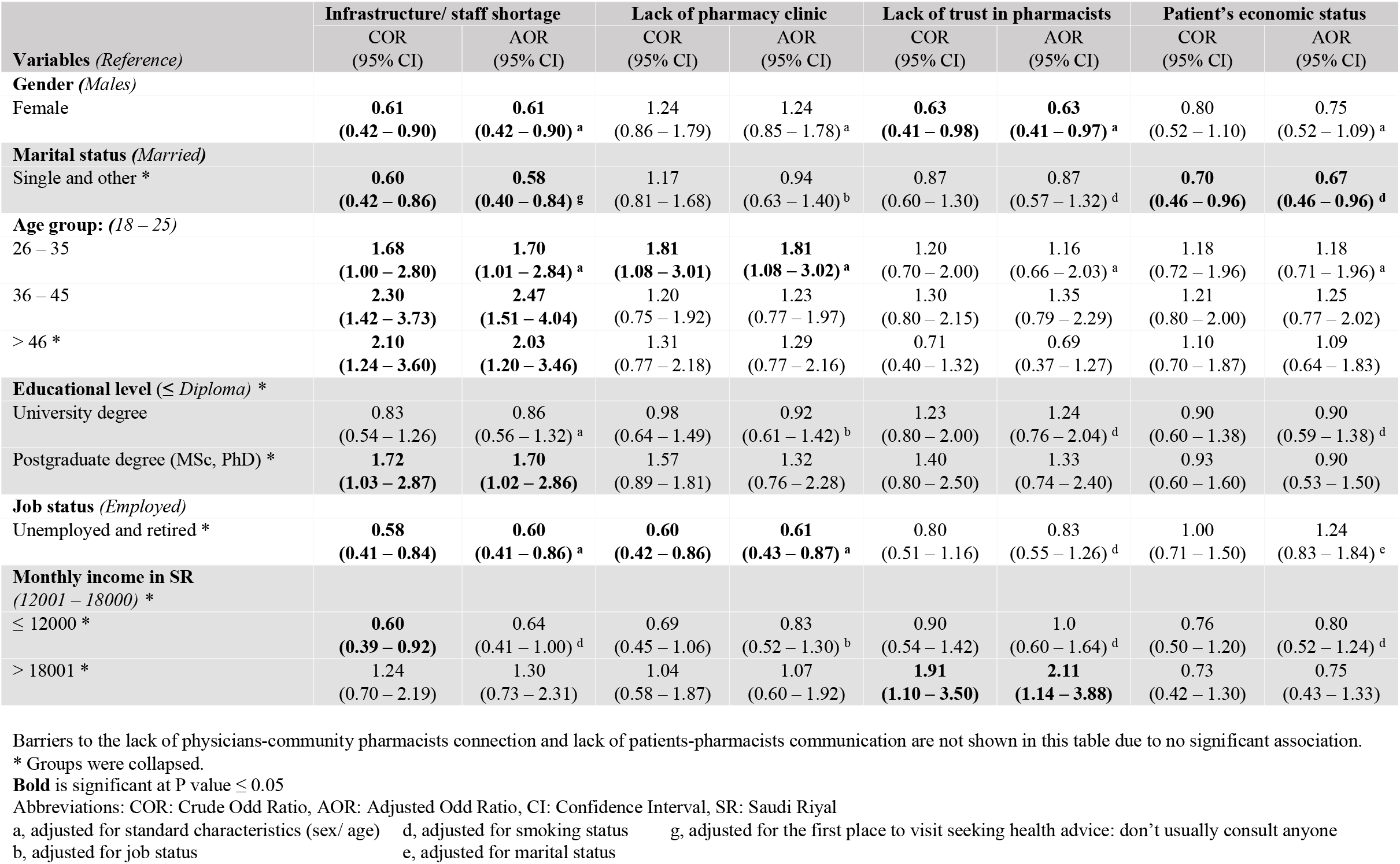
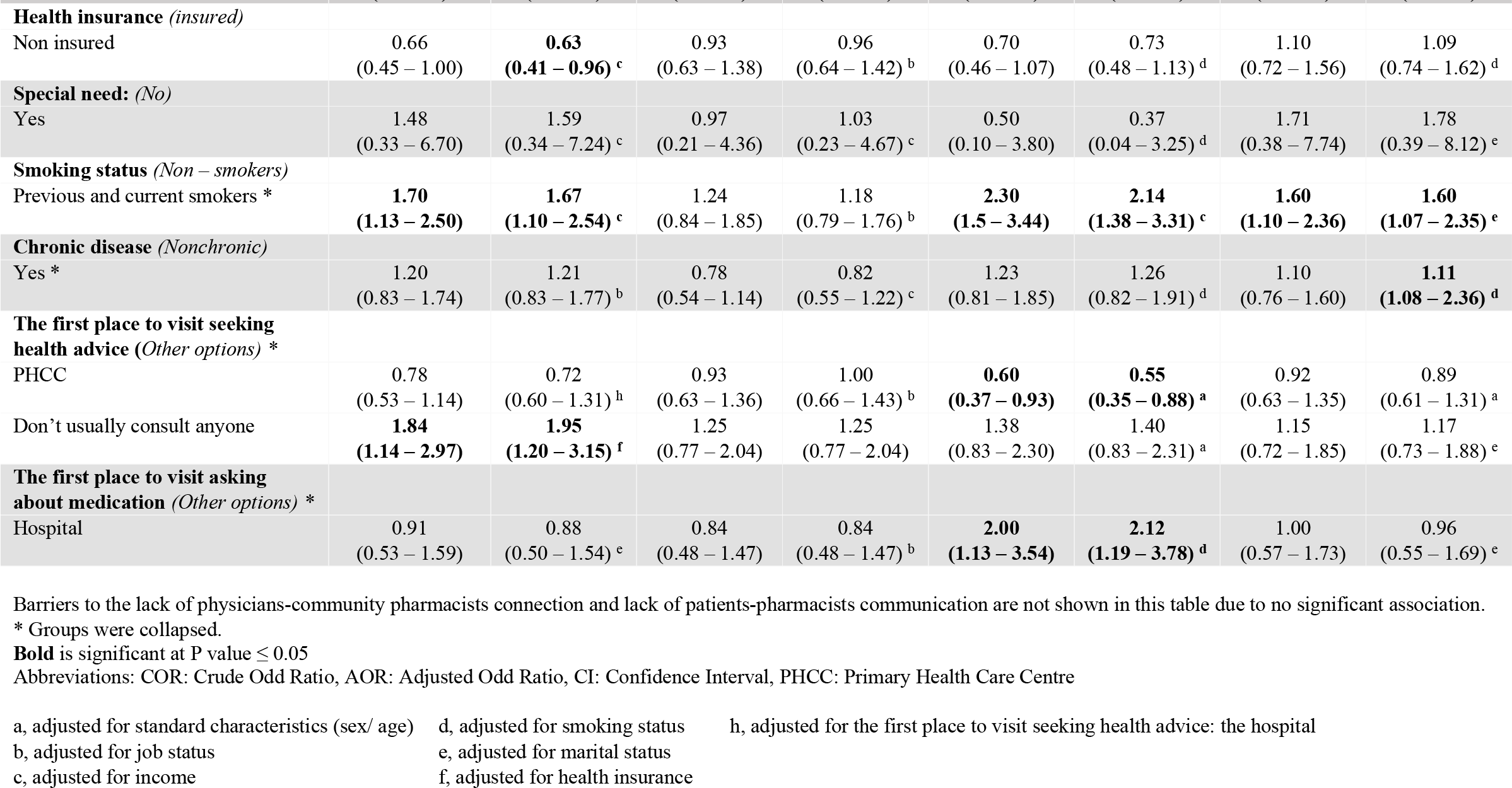
Crude and adjusted logistic regression of the association between barriers to using ECPSs and characteristics of participants in Saudi Arabia. N = 533.

## Discussion

The world has observed a significant transition in the pharmaceutical care concept from its traditional model, which is merely concerned with dispensing practice, to a more interprofessional model with broader but crucial public health objectives. In 2030, SA will be welcoming two major events with national and global agendas. The first is the assessment of the Sustainable Development Goals (SDGs), where target 3.4, “to reduce by one-third premature mortality from non-communicable diseases through prevention and treatment” [25], will be assessed. The second major event is completing and evaluating the Saudi 2030 national vision projects [26]. Both events have played a significant role in driving the health system re-engineering process currently underway in SA. Hence, changes anticipated for community pharmacy practices will essentially act as substantial factors from the public’s perspectives as healthcare receivers of ECPSs and can be translated into a practical pharmaceutical care approach.

In this study, the respondents were all CPs customers, including patients. Almost one-third considered CPs the first destination to visit for seeking health advice when a minor ailment arises. Published reports from various other sources demonstrated a broad range of this pattern, such as in Ethiopia (30%) [8], Portugal (36%) [19], Malaysia (46%) [27], Kuwait (61%) [19], and Qatar (91%) [12], and this may reflect different social, health systems and economic aspects. Despite the high access rate to the PHCC in SA due to free of charge health system model, the role of CPs in providing timely, easy, and relatively cheap medical advice can support the country’s medical services in responding to more severe health conditions.

Another prime reason for the public to visit CPs was collecting POM and OTC, almost half of the participants were either practising self-medication or collecting prescribed medication by physicians via CPs. This general tendency is consistent with findings from other national studies [28-30]. Notably, only 12% of the study respondents’ purpose for visiting CPs was seeking other healthcare services such as ECPSs. That only minimal services could explain this small proportion of interest eventually reaching the public, which can be supported by the observation of low experience among respondents about ECPSs, as revealed in this study.

Moreover, Most respondents needed to know that CPs can provide medication therapy management, health screening, bodyweight management, smoking cessation services and others. The lack of ECPSs in the local CPs is probably due to three key factors: a) high standards and requirements set by the concerned Saudi authorities against ECPSs implantation, b) low functionality or professionality of CPs, or c) low public awareness due to low priority for integrating the introduction to ECPSs in wider public communication platforms. The latter factor is similar to other settings as reported elsewhere; although ECPSs have existed in the United Kingdom since 2005, the public still needs to be aware of the full role of the CPs [31].

During the COVID-19 pandemic, the Saudi MOH signed a memorandum with one of Saudi largest CPs chains to deliver COVID-19 vaccines [32], but it was only limited to large provinces in SA. The capacity and readiness of CPs in these provinces for the assigned services are probably more than those in small provinces. Raising awareness of public health aspects among people is a crucial initiative by governments. This may explain the high proportion of public knowledge of the availably of immunisation services in Saudi CPs.

The Saudi MOH has launched contact channels with the public. Moreover, a free 24-hour call centre was introduced in 2016 to receive public concerns regarding any health issue, followed by the e-health application called “Sehhaty” to facilitate visual medical consultations on smartphones [33, 34]. However, results showed a deficient proportion of using these applications. The participants tend to search the internet websites more than contacting the MOH channels. Long queues while using these channels or low public awareness may explain this low proportion.

In 2018, the Saudi MOH released, through multiple stages, the Wasfaty system using a national electronic prescription platform through only contracted CPs [35]. Nevertheless, nearly half of the participants needed to be aware of this platform. Additionally, electronic medical records are inaccessible to local CPs. Still, two-thirds of the public has yet to experience this facility before. A study by Al Hassan et al. in 2010 called for developing guidelines or regulations for professional practice relating to patients’ medical records [36]. However, this facility has only been available in a few other countries [37]. Linking patient medical history to CPs through a regulated platform would efficiently facilitate a modern digitalised form of enhanced physician-pharmacist-patient communication, which can have significant implications for public health and healthcare delivery.

The study results show that the public strongly supports ECPSs implementation in CPs with a high degree of importance, a positive attitude, and a desire with the potential to support future scaling up of such services. This attitude is consistent with several global studies [6, 14, 15, 31, 38]. Nevertheless, there are several barriers and public concerns towards these services.

Against the direct assessment of the SDGs target 3.8, “achieving universal healthcare access” [39], barriers to using ECPSs have been processed through in-depth logistic regression analysis. The CPs infrastructure, including the healthcare workforce allocated for these services, has been important in the public’s opinion. Almost all respondents’ background and characteristics factors (socio-demography, economy, health status and behaviours) have been associated with the CPs underdeveloped infrastructure and staff shortages as a key barrier.

Nevertheless, the lack of pharmacy clinics to deliver private counselling was significant, as declared in this study, where most respondents did not receive confidential counselling inside CPs. This finding is consistent with previous national studies highlighting public privacy and confidentiality concerns [40-44]. A systematic international review of feedback from CPs users by Claire Anderson et al., 2004 considered privacy and confidentiality when providing advice as critical factors to increase the acceptability of CPs’ roles [45]. Furthermore, male community pharmacists dominate the private sector in SA (78%), and the proportion can increase in CPs settings [46]. This corresponds to the higher probability of having females being risk factors for the lack of privacy due to the unavailability of pharmacy clinics. This is also addressed elsewhere, where 58% of Saudi females favour counselling by female pharmacists [47]. In contrast, participants with different backgrounds also have concerns regarding their privacy in this study. Therefore, Saudi CPs must be better designed and equipped with good facilities for a safe and pleasant environment where pharmacist-patients exchange sensitive information.

The respondents’ economic status has been an additional barrier associated with several variables in this study since two-thirds disagreed or were neutral if any ECPSs are being paid services, particularly where only 25% of the public possess health insurance. Securing health insurance enhances the population to seek medical check-ups in the Saudi private health sector [48]. In SA, all private sector employees and their family members are health insured by law. During this study, a new Saudi law has been introduced, which grants government employees and their families full health insurance that also covers healthcare services in the private sector. However, not all government agencies grant their employees with health insurance. Therefore, health insurance coverage remains limited to agencies permitted by labour regulation [49]. This economic barrier should be considered essential when there is no universal health insurance coverage.

Although 73% of study subjects showed a general trust in community pharmacists, a lack of patient-community pharmacist communication was observed in this study. More than half of respondents consider it a potential barrier. Moreover, workload and staff shortage in the absence of scheduled appointments in the CPs can lead to poor communication with the community pharmacists. In comparison, the lack of inter-professional connections between physicians and community pharmacists to discuss patients’ health conditions was considered by the participants. Evidence where physician-pharmacist collaboration in clinical settings has shown to improve public health outcomes [38]. However, there is a lack of social and professional acceptance of community pharmacists among physicians in SA [50]. Such factors affect the overall pharmaceutical care process.

## Conclusion

This study is timely concerning the upcoming national and global agendas and will likely provide valuable findings to guide future policies. As this study shows, ECPSs and CPs facilities need to be adequately utilised by the public despite their appreciation of services implementation. Fundamental aspects perceived by the public related to their privacy, the cost of services, and lack of professional communication where such ECPSs are provided, whereas some groups are at higher risk of being exposed to these barriers. In addition, CPs must be better designed and equipped with facilities for good pharmacy practice. Finally, it is worth noting that the Wasafaty system by MOH and online pharmaceutical care, including prescription delivery by CPs, are relatively new concepts in SA. Therefore, they require further research and regular evaluation to assess their application and progression.

### Limitations

The choices for the sampling and study design of this study have been affected by the strict COVID-19 pandemic distancing regulations. Relying on a self-administered web-based questionnaire as a source of information in cross-sectional design can raise some concerns about how findings can be interpreted and generalised. Nevertheless, despite its non-probability nature, the integration of the snowball sampling approach has enhanced the response rate and added value to the randomness of questionnaire distribution via web applications. In addition, under-representative samples were observed in this study across elderly participants, females, people with associated chronic diseases, people with special needs and health insurance. Furthermore, some responses were skewed, as noted in a few outcome categories. However, the descriptive nature of the study design coupled with a careful and thorough multiple regression process can adjust associated issues attributed to the imbalanced distributions of some groups.

## Data Availability

All relevant data are within the manuscript and its Supporting Information files.

## Author Contributions

**Conceptualisation:** Khalid S. Alghamdi, Max Petzold, Laith Hussain-Alkhateeb.

**Data curation:** Khalid S. Alghamdi, Max Petzold, Ashraf A. Ewis, Mahdi H. Alsugoor, Laith Hussain-Alkhateeb.

**Formal analysis:** Khalid S. Alghamdi, Max Petzold, Ashraf A. Ewis, Laith Hussain-Alkhateeb.

**Investigation:** Khalid S. Alghamdi, Laith Hussain-Alkhateeb.

**Methodology:** Khalid S. Alghamdi, Max Petzold, Laith Hussain-Alkhateeb.

**Project Administration:** Max Petzold, Laith Hussain-Alkhateeb

**Supervision:** Max Petzold, Laith Hussain-Alkhateeb.

**Validation:** Ashraf A. Ewis, Khalid Saaban, Mahdi H. Alsugoor.

**Visualisation:** Khalid S. Alghamdi

**Writing – original draft:** Khalid S. Alghamdi, Laith Hussain-Alkhateeb

**Writing – review and editing:** Ashraf A. Ewis, Max Petzold, Khalid Saaban, Mahdi H. Alsugoor, Laith Hussain-Alkhateeb.

## Acknowledgements

The authors sincerely appreciate the effort by Miss. Wejdan Alghamdi, Linguistics MA student, UCL, UK., for the translation of the questionnaire into Arabic language. The authors extend their appreciation to Mrs Amal Alghamdi, Ministry of Education, Riyadh, SA, for revising the Arabic questionnaire and grammatical corrections.

## Supporting information

S1 File. Public questionnaire (English and Arabic Versions)

S2 File. Data

## Notes

### Competing Interest Statement

The authors have declared that no competing interests exist.

### Funding Statement

This study is a part of a PhD research project sponsored by Al Baha University, Saudi Arabia (Decision of Scholarship No: 40267573, July 2019).

### Author Declarations

Permission for conducting the study was obtained from the Ethical Research Committee at the Saudi Ministry of Health (MOH), Riyadh (Central IRB log No: 21-56E, June 2021).

